# Integrating anamnestic and lifestyle data with sphingolipid levels for risk-based prostate cancer screening

**DOI:** 10.1101/2025.03.07.25323552

**Authors:** Caterina Peraldo-Neia, Paola Ostano, Melissa Savioli, Maurizia Mello-Grand, Ilaria Gregnanin, Francesca Guana, Francesca Crivelli, Francesco Montagnani, Michele Dei-Cas, Rita Paroni, Antonella Sinopoli, Francesco Ferranti, Nicolò Testino, Marco Oderda, Andrea Zitella, Chiara Fiameni, Amedeo Gagliardi, Alessio Naccarati, Luca Clivio, Paolo Gontero, Stefano Zaramella, Giovanna Chiorino

## Abstract

**Background:** In the era of risk-based prostate cancer (PCa) screening, overcoming the limitations of prostate-specific antigen (PSA) testing and stratifying men by individual risk is crucial. Our study aims to integrate anamnestic and lifestyle data with circulating biomarkers to minimize unnecessary second-level investigations (SLIs) for patients with suspected PCa, while improving the detection of clinically significant PCa (ISUP>1).

**Methods:** We collected plasma samples, recent clinical history, family cancer history, PSA levels, and lifestyle information from 904 men: 421 undergoing PSA testing, 421 with suspected and 62 with confirmed PCa. Univariable logistic regression was applied to identify ananmestic and lifestyle variables mostly associated with PCa. Penalized logistic regression models predictive of PCa or ISUP>1 PCa were built both using the 814 subjects with complete information for such variables, applying a 10-fold cross validation approach, and dividing the dataset into a training (n=445: 132 PCa, 313 non-PCa) and a test (n=369: 147 PCa, 222 non-PCa) set. The concentration of 50 sphingolipids was analysed on the latter set of 369 subjects by mass-spectrometry, and multivariable penalized regression with 10-fold cross-validation was applied to integrate anamnestic, lifestyle, sphingolipid data. ROC-AUCs on the test sets were compared with PSA ROC-AUCs.

**Results:** Age, cardiovascular disease (CVD), number of medications, and sedentariness were significantly associated with PCa detection and their combination with PSA improved its performance (ROC-AUC from 0.85 to 0.89). In the SLI subgroup (n=437), adding age improved PSA predictive power (ROC-AUC from 0.60 to 0.70), but performance was still poor. Penalized regression with 10-fold cross-validation on the sphingolipid dataset identified hypertension, CVD, PSA, age, and five sphingolipids (HexCer-20, Cer-20, HexCer-24.1, GM3-24.1, DHCer-24) as key variables for accurate PCa classification (average ROC-AUC: 0.92). Cer-20 and CVD were consistently selected by models predicting ISUP>1 PCa. In the SLI subgroup, PSA, age, CVD, SM-16, HexCer-20, HexCer-24.1, DHS1P, and DHCer-24 were selected in all 10 models (average ROC-AUC: 0.83).

**Conclusions:** Circulating sphingolipids are promising biomarkers that, when combined with PSA, anamnestic, and lifestyle data, may enhance PCa screening precision and reduce the need for invasive, costly examinations.

## Introduction

Prostate cancer (PCa) is the most commonly diagnosed cancer and ranks as the fifth leading cause of cancer-related deaths among men in Italy (https://gco.iarc.who.int/en; Globocan 2022, version 1.1). Digital rectal examination (DRE) has low sensitivity in identifying PCa [1]. Measuring ematic prostate-specific antigen (PSA) levels enables early PCa detection, but presents some limitations: aggressive tumors can be missed in men with PSA levels below 3 ng/ml and higher PSA levels do not automatically correspond to a PCa diagnosis. This last aspect leads to the significant downside of unnecessary invasive biopsies for a large fraction of men [2]. Multiparametric magnetic resonance imaging (MRI) offers high accuracy in detecting clinically significant PCa (csPCa), defined by International Society of Urological Pathology (ISUP) grade higher than 1 (ISUP>1), therefore reducing the need for unnecessary biopsies. However, it is resource-intensive, requiring experienced radiologists, considerable time, and significant costs. A set of biomarkers able to complement PSA and DRE in the identification of patients who require further clinical examinations is mandatory to spare patients without PCa from unpleasant, costly, and invasive procedures.

Starting from the analysis of plasma samples collected before diagnostic biopsy from nearly 400 men, we previously uncovered a possible role of ceramides in discriminating PCa from benign prostatic hyperplasia when PSA was in the so-called “grey zone”, with 40% reduction of PSA false positives [3]. Ceramides are structurally defined as sphingoid bases, typically sphingosine with 18 carbons, attached to a fatty acyl chain of variable length (14 to 26 carbons), the most common one being 16 carbons in mammalian cells [4]. Ceramides are essential intermediates in the biosynthesis and metabolism of all sphingolipids and regulate many pathways, such as apoptosis, cell cycle, cell senescence and differentiation, through the synthesis and degradation of different sphingolipids [5]. Their homeostasis is crucial and their role in regulating biological processes is still partly unexplored, with some species associated with a tumor suppression and other to a tumor promoting function [6]. Interestingly, aberrations in circulating species of the sphingolipid pathways have been consistently found in patients with localized PCa that clinically progressed or with metastatic/castration therapy resistant PCa [7–9], in PCa resistant to docetaxel [10] or to androgen receptor signalling inhibitors [11], as well as in neuroendocrine PCa [12] and in prostate cells treated with fatty acid synthase inhibitor, highlighting a role also in mediation of lipid damage [13].

Recent advancements in metabolomic profiling have evidenced a dysregulation of amino acids and sphingolipid metabolism during the development of PCa [14]. Case-control studies from the European Prospective Investigation into Cancer and Nutrition (EPIC) cohort [15–17] have highlighted patterns of circulating metabolites, including sphingolipids, associated with PCa cancer risk or with the development of aggressive PCa. However, to our knowledge no study has yet exploited samples from men undergoing longitudinal PCa screening or diagnostic biopsy to measure such molecules, alone or in combination with PSA and individual risk factors.

In this work, we took advantage of a prospective collection of plasma samples and questionnaire records from men undergoing organized PSA testing and DRE, prostate biopsy for suspected PCa, or radical prostatectomy. Information on lifestyle, cancer familiarity and comorbidities was analyzed in association with PCa diagnosis, to highlight the most important risk factors, which were then combined with PSA, and circulating sphingolipid levels. Penalized regression models were always developed and tested 10 times, using a 10-fold cross-validation approach, to avoid overfitting. We aimed at: i) better selecting men who need second level investigations (SLI), to avoid unnecessary exams; ii) discriminating men with csPCa or high grade PCa from the rest, to avoid overdiagnosis or overtreatment.

## Material and methods

### Cohort recruitment, sample and data collection

421 men (age range: 50-69) undergoing opportunistic PSA screening at Fondazione Edo ed Elvo Tempia (FEET) were prospectively recruited between September 2021 and October 2023. Blood sampling was carried out at recruitment and every six months, for at least three times. All the recruited subjects were checked by a urologist and underwent DRE. Those recalled for SLI were followed-up according to the European Association of Urology (EAU) guidelines (https://uroweb.org/guidelines/prostate-cancer). Sixty-two PCa patients were prospectively recruited at the Biella Hospital (BH) between October 2022 and February 2024. Blood sampling was carried out before surgery and at each follow-up, according to the EAU guidelines. 421 men were prospectively recruited at Molinette Hospital (MH) before diagnostic biopsy for suspected PCa, and blood sampling was carried out at recruitment. Subjects with negative DRE and no alteration of PSA for at least 4 consecutive evaluations were named healthy donors, “HD”; these, together with subjects with negative MRI/biopsy, were named “nonPCa”.

Self-reported questionnaires on lifestyle (physical activity, body mass index (BMI), diet, smoke and alcohol consumption), familiarity, past prostate biopsies and concurrent pathologies, as well as PSA levels and DRE outcomes, have been collected for each subject recruited at FEET, BH and MH. An online password protected database (https://www.keyform.eu) was used to upload the electronic Case Report Forms (eCRF) including questionnaires, urological visit report, PSA dosage and lab analyses at each follow-up, as well as histological and clinical information for PCa. Each patient was pseudoanonymized and signed an informed consent.

The observational clinical protocol called DP3 coordinated by FEET was approved by the Novara ethical committee (AOU Maggiore della Carità) in September 2020 (Prot. N° 968/CE 07/09/2020, integration Prot. N°263/CE, 10/03/2021), in accordance with the Declaration of Helsinki Ethical Principles for Medical Research involving Human Participants. The MH ethical committee (AOU Città della Salute e della Scienza) in Turin approved the DP3 study in January 2022 (Prot. N° 521/2021, 10/01/2022).

A summary of the cohorts available is depicted in **Table 1**.

**Table 1.** Site distribution of patients/samples available, tumor characteristics and analyses performed.

| **Sites** | **FEET** | **BH** | **MH** | **total** |
| --- | --- | --- | --- | --- |
| Total recruited (Blood sampling at enrolment – Plasma isolation and storage – PSA – DRE) | 421 | 62 | 421 | **904** |
| Blood sampling every 6 months for at least 4 times | 408 | na | na | **408** |
| Blood sampling after surgery as per clinical practice | 6 | 62 | na | **68** |
| Questionnaire filled | 421 | 62 | 421 | **904** |
| Complete data for PSA, age, hypertension, CVD, PA level, medications | 417 | 60 | 337 | **814** |
| Targeted sphingolipid analysis | 182 | 56 | 155 | **393** |
| Recalled for SLI | 40 | 62 | 421 | **523** |
| PCa diagnosed | 13 | 62 | 260 | **335** |
| ISUP>1 PCa | 6 | 56 | 234 | **296** |
| ISUP >2 PCa | 2 | 37 | 143 | **182** |

FEET: Fondazione Edo ed Elvo Tempia; BH: Biella Hospital; MH: Molinette Hospital; SLI: Second level investigation; CVD: cardiovascular diseases; PA: physical activity

### Blood sample processing

Whole blood was collected in 3 EDTA K2 tubes (10 ml each, D.P. MEDICAL s.r.l., Centallo (TO), Italy) by venipuncture. Plasma was isolated within 1 hour from blood sampling. FEET and BH samples were processed at the Genomics Lab of FEET while MH samples were processed at MH. For plasma separation, tubes were centrifuged at 2500 rpm for 10 min at 4°C; supernatant was then collected in new 15 ml tubes and centrifuged at 2500 rpm for 10 min at 4°C again; plasma was then aliquoted in barcoded tubes and stored at -80°C. Frozen plasma samples from MH were then sent to the Genomics Lab of FEET.

### PSA evaluation

PSA concentration was centrally evaluated at BH for all the men recruited at FEET and BH using Elecsys total PSA kit IVD (Cobas, Roche Diagnostics, Monza, Italy). For the men recruited at MH, pre-biopsy PSA records were shared with FEET, according to the study protocol and the data transfer agreement.

### Targeted sphingolipid analysis and processing

Frozen plasma aliquots were sent to the Department of Health Sciences, University of Milan, where 50 species of the sphingolipid metabolism (Ceramides, dihydroCeramides, sphingomyelins, hexosylCeramides, lactosylCeramides, globotriaosylCeramides, gangliosides GM3, sphingosines – see complete list in **Additional file 1**) have been analyzed in a subset of 369 plasma samples by mass spectrometry. Briefly, from 25 µL of plasma, sphingolipids were isolated by monophasic extraction (methanol/chloroform 2:1, Merk Life Science S.r.l., Milan, Italy) coupled to alkaline methanolysis. The extracts were resuspended in pure methanol and injected in a LC-MS/MS apparatus: Dionex 3000 UPLC, (ThermoFisher Scientific, Monza, (MB), Italy), coupled to 3200 QTRAP, (AB Sciex S.r.l. Milan, Italy). The chromatographic separation was attained on a C8 column for complex sphingolipids and a C18 column for sphingoid bases.

Four species were undetectable in most of the samples and 46 sphingolipids were retained for the analysis. Z-score standardization was performed on raw data, and batch effect was corrected using the ComBat function from the *sva* package in R.

### Lifestyle score calculation

A lifestyle score reflecting concordance with the World Cancer Research Fund (WCRF) Recommendations [18] and the indications of the Italian Guidelines for a healthy diet [19] was defined. Starting from questionnaire data available, five parameters were considered: BMI, physical activity, dietary habits, smoking habit and alcohol consumption. A value between -1 and +1, based on a lesser or greater adherence to the WCRF Recommendations and the indications of the Italian Guidelines, was assigned to each parameter, as detailed below. For BMI score, the value of 1 was given to each subject whose BMI was in the range of healthy weight (18.5-24.9), 0.5 for underweight (< 18.5), 0 for overweight (25.0-29.9), -1 for obesity (≥ 30). For physical activity (PA), self-reported engagement in high level PA (3 or more times a week) was assigned a value of 1, moderate PA (1-2 times a week) a value of 0, while sedentariness a value of -1. For smoking, 1 was assigned to those subjects who reported not smoking or quitting more than 15 years ago, 0 to those who had quit less than 15 years prior and -1 to current smokers. For alcohol, 1 was given for no consumption, 0.5 was given for alcohol consumption frequency of 1-2 times a week, 0 for a frequency of 3-4 times a week and -1 for a daily alcohol consumption. For diet, the definition of a score was more complex because of the variety of food items and the requirement to assess the overall dietary pattern of each subject. Calculation details are reported in the **Additional file 2**.

### Statistical analyses

The Shapiro-Wilk normality test was applied to continuous variables in order to test the normality of their distributions. Log transformation was applied when the normality assumption was not satisfied. Univariable and penalized multivariable logistic regression analysis were applied to identify features associated with PCa diagnosis, using the entire cohort. Samples with complete information on the mostly assocated variables were retained for subsequent analyses. On the resulting samples, two approaches (step1) were followed (**Figure 1**): Step 1a) least absolute shrinkage and selection operator (LASSO) penalized logistic regression, with 10-fold cross-validation, was applied to retrieve the variables selected by at least 9/10 models (in each dataset partition, the same proportion of cases and non-cases as in the original dataset was maintained); Step 1b) the entire cohort was divided into a training test used to build a LASSO model, which was then applied on a test set. In Step 2, sphingolipid levels, available only for the test set, were included in the analysis and combined with anamnestic/lifestyle variables by LASSO regression with the 10-fold cross-validation approach. Area under the Receiver Operator Curve (ROC AUC) was used to assess the discriminatory ability of single variables or logistic regression models, with reported 95% confidence intervals (C.I.). Paired DeLong test was used to compare the discrimination among different variables/models and Student paired t-test to compare the average AUCs between the LASSO models and PSA or PSA plus age.

**Figure 1:**
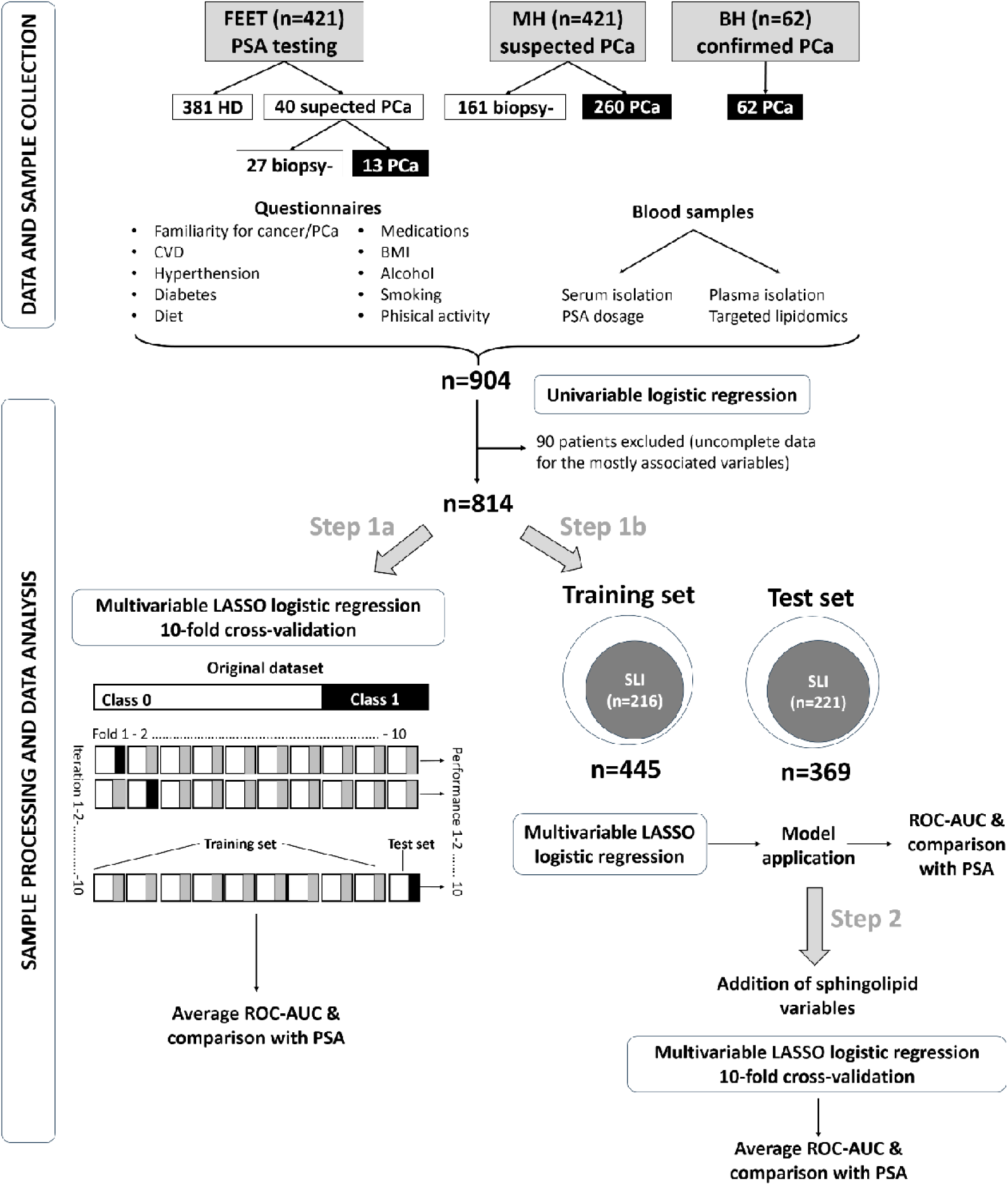
Workflow of the study. FEET: Fondazione Edo ed Elvo Tempia; BH: Biella Hospital; MH: Molinette Hospital; HD: Healthy donors; CVD: cardiovascular diseases; BMI: Body Mass Index. Black rectangulars: PCa. White rectangulars: non PCa.

Wilcoxon and Chi-square tests were applied to compare continuous and categorical covariates between groups, respectively.

All analyses were performed using the *glmnet*, *pROC*, *ggpubr*, and *stats* packages within the statistical computing environment R (version 4.2.1). Associations in the univariable logistic regression analysis were considered statistically significant if the p-value was below 0.001. Differences were considered statistically significant if the p-value of the Wilcoxon, Chi-square, DeLong or Student t-tests were below 0.01.

## Results

### Sample characteristics

904 plasma samples from men recruited in the DP3 study were collected at the baseline (**Table 1**).

Within the MH cohort, 260 out of 421 subjects (61.8%) had a positive biopsy. Among men recruited at FEET for opportunistic PSA screening, 40 subjects underwent SLI and 13 out of 40 (32.5%) were classified as PCa: three are under active surveillance, one was treated with radiotherapy, three had prostatectomy in other hospitals than BH, six were surgically treated at BH and were followed-up as per clinical practice, like the 62 PCa patients from BH. The study workflow is depicted in **Figure 1** and information obtained from the questionnaire completed at enrolment is reported in **Table 2**.

**Table 2:** Characteristics of the men at the enrolment based on questionnaire responses.

|  | Entire cohort | Non-PCa | PCa | P-value |
| --- | --- | --- | --- | --- |
| Age (median, range),<br>n=902 | 64.7<br>(43.2-88.9) | 61.7<br>(48.96-82.6) | 69.7<br>(43.2-88.9) | <0.00001 |
| PSA (median, range),<br>n=903 | 4.363<br>(0.02-1000) | 1.96<br>(0.02-39.4) | 7<br>(0.5-1000 ) | <0.00001 |
| Familiarity, cancer (%),<br>n=837 | 53.4 | 52.6 | 54.9 | 0.5662 |
| Familiarity, PCa (%),<br>n=823 | 12.6 | 11.3 | 15.2 | 0.1367 |
| Cardiovascular diseases<br>(%), n=875 | 13.6 | 9.1 | 21.5 | <0.00001 |
| Diabetes (%), n=882 | 8.05 | 6.4 | 10.9 | 0.0259 |
| Hypertension (%), n=881 | 44.6 | 37.97 | 56.3 | <0.00001 |
| Medications (%), n=880 | 67.3 | 60.4 | 79.4 | <0.00001 |
| BMI (median, range),<br>n=832 | 25.6<br>(14.9-48.2) | 25.4<br>(14.9-48.2) | 26.1<br>(19.3-37.0) | 0.0439 |
| Physical activity<sup>1</sup> (%),<br>n=833 | 26.9 | 22.9 | 34.4 | 0.000497 |
| Smoke<sup>2</sup> (%), n=848 | 25.6 | 26.3 | 24.3 | 0.5906 |
| Alcohol<sup>3</sup> (%), n=833 | 71.7 | 75.3 | 64.8 | 0.0018 |

<sup>1</sup>baseline for physical activity: sedentary; <sup>2</sup> smoker or previous smoker who quit less than 15 years before enrolment; <sup>3</sup> consumers, with any frequency.

### Lifestyle and risk factors

All the 904 enrolled men within the DP3 cohorts received a questionnaire on lifestyle, familiarity, past biopsies and concurrent pathologies. The forest plot for the main clinical features is reported in **Figure 2**.

**Figure 2:**
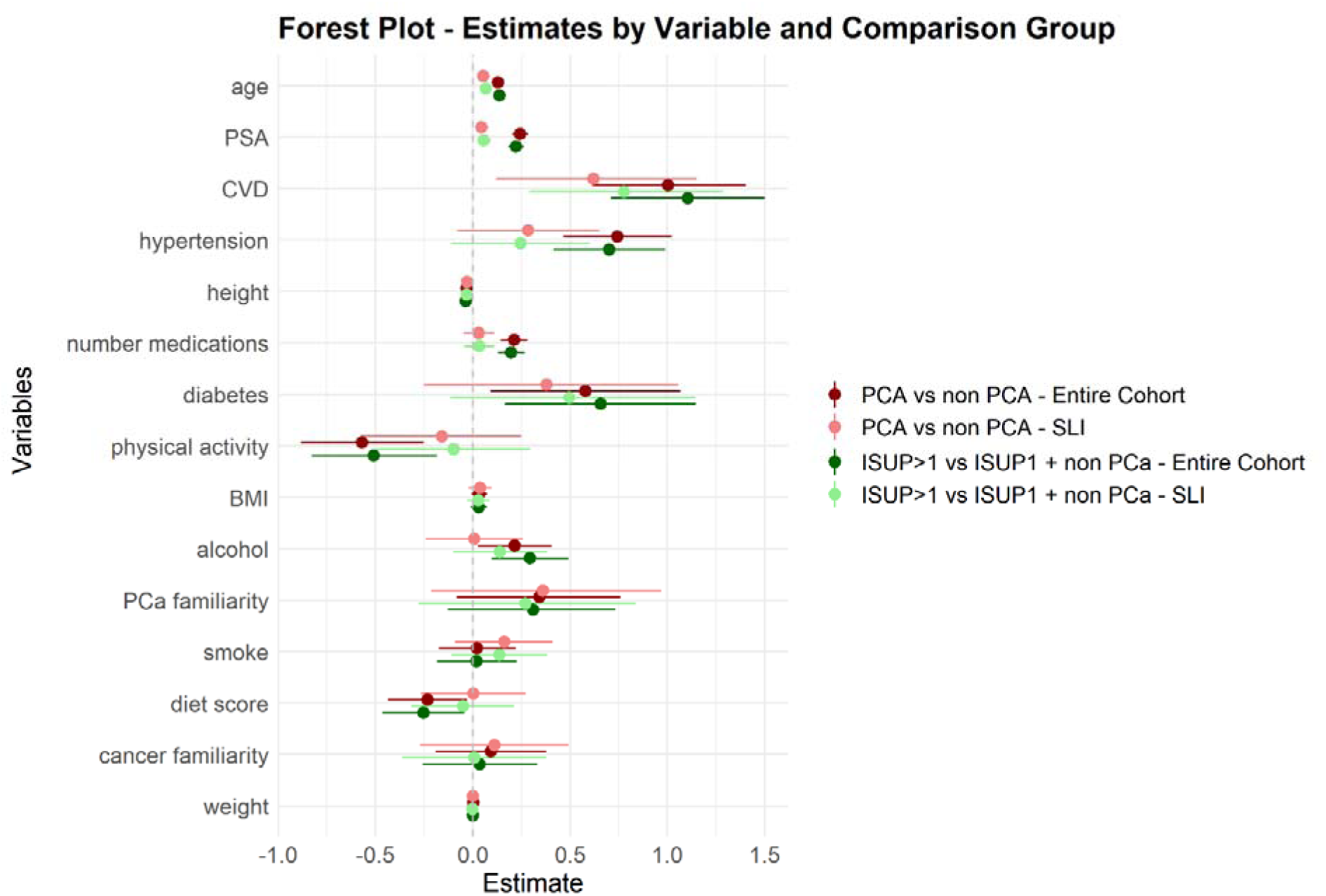
Forest plot of clinical variables comparing PCa vs nonPCa (dark red for analyses on the entire cohort, pink for SLI) or ISUP>1 PCa vs ISUP1_Pca + nonPCa (dark green for analyses on the entire cohort, light green for SLI).

The analysis of questionnaires revealed that age, pharmacological treatments in general, hypertension, cardiovascular diseases (CVD) and physical activity (PA) were significantly associated with PCa, independently of PSA. Age, CVD and the number of medications were positively associated also with ISUP>1 PCa when ISUP1 tumors were included in the reference group, independently of PSA (**Table 3**).

**Table 3:**
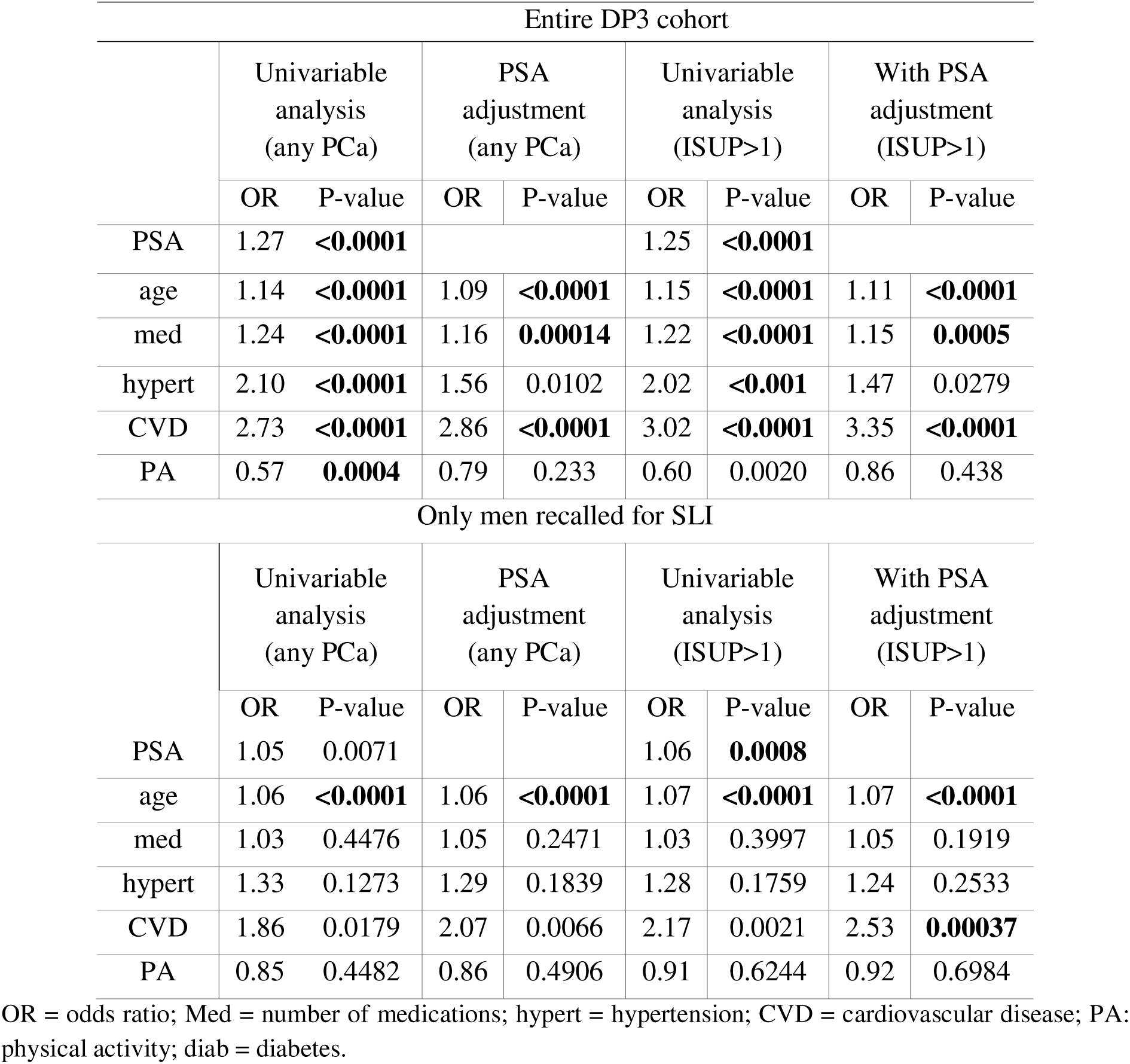
Univariable or multivariable (with PSA adjustment) logistic regression of selected variables.

| Entire DP3 cohort |  |  |  |  |  |  |  |  |
| --- | --- | --- | --- | --- | --- | --- | --- | --- |
|  | Univariable analysis (any PCa) |  | PSA adjustment (any PCa) |  | Univariable analysis (ISUP>1) |  | With PSA adjustment (ISUP>1) |  |
|  | OR | P-value | OR | P-value | OR | P-value | OR | P-value |
| PSA | 1.27 | **<0.0001** |  |  | 1.25 | **<0.0001** |  |  |
| age | 1.14 | **<0.0001** | 1.09 | **<0.0001** | 1.15 | **<0.0001** | 1.11 | **<0.0001** |
| med | 1.24 | **<0.0001** | 1.16 | **0.00014** | 1.22 | **<0.0001** | 1.15 | **0.0005** |
| hypert | 2.10 | **<0.0001** | 1.56 | 0.0102 | 2.02 | **<0.001** | 1.47 | 0.0279 |
| CVD | 2.73 | **<0.0001** | 2.86 | **<0.0001** | 3.02 | **<0.0001** | 3.35 | **<0.0001** |
| PA | 0.57 | **0.0004** | 0.79 | 0.233 | 0.60 | 0.0020 | 0.86 | 0.438 |
| Only men recalled for SLI |  |  |  |  |  |  |  |  |
|  | Univariable analysis (any PCa) |  | PSA adjustment (any PCa) |  | Univariable analysis (ISUP>1) |  | With PSA adjustment (ISUP>1) |  |
|  | OR | P-value | OR | P-value | OR | P-value | OR | P-value |
| PSA | 1.05 | 0.0071 |  |  | 1.06 | **0.0008** |  |  |
| age | 1.06 | **<0.0001** | 1.06 | **<0.0001** | 1.07 | **<0.0001** | 1.07 | **<0.0001** |
| med | 1.03 | 0.4476 | 1.05 | 0.2471 | 1.03 | 0.3997 | 1.05 | 0.1919 |
| hypert | 1.33 | 0.1273 | 1.29 | 0.1839 | 1.28 | 0.1759 | 1.24 | 0.2533 |
| CVD | 1.86 | 0.0179 | 2.07 | 0.0066 | 2.17 | 0.0021 | 2.53 | **0.00037** |
| PA | 0.85 | 0.4482 | 0.86 | 0.4906 | 0.91 | 0.6244 | 0.92 | 0.6984 |

OR = odds ratio; Med = number of medications; hypert = hypertension; CVD = cardiovascular disease; PA: physical activity; diab = diabetes.

The final diet score was negatively associated with PCa, indicating a protective role, even if with low statistical significance (OR=0.80; p-value=0.035). As expected, PSA was significantly associated with PCa in the entire prospective cohort (**Table 3**).

To test whether a combination of the clinical covariates with significant association to PCa could improve the discriminatory ability of PSA and age, we considered the subset with complete information on PSA, age, hypertension, CVD, sedentariness and the number of medications (n=814). We first applied penalized LASSO logistic regression modelling with 10-fold cross-validation (**Figure 1**, step 1a) and obtained 10 models, tested each time on the 1/10 of samples not used for model construction. The average ROC AUC on the 10 test sets was 0.89 (range: 0.84-0.93), while for PSA it was 0.85 (range: 0.81-0.91) (**Figure 3A**, **left panel**). PSA, age, hypertension and CVD were selected by 10/10 models and PA by 9/10, indicating their relevance for the identification of men who really need SLI and for the implementation of targeted prevention actions. When we considered only the men who underwent SLI (n=437), the LASSO models did not select any anamnstic/lifestyle variable and showed poor performance (**Figure 3B, left panel**).

**Figure 3:**
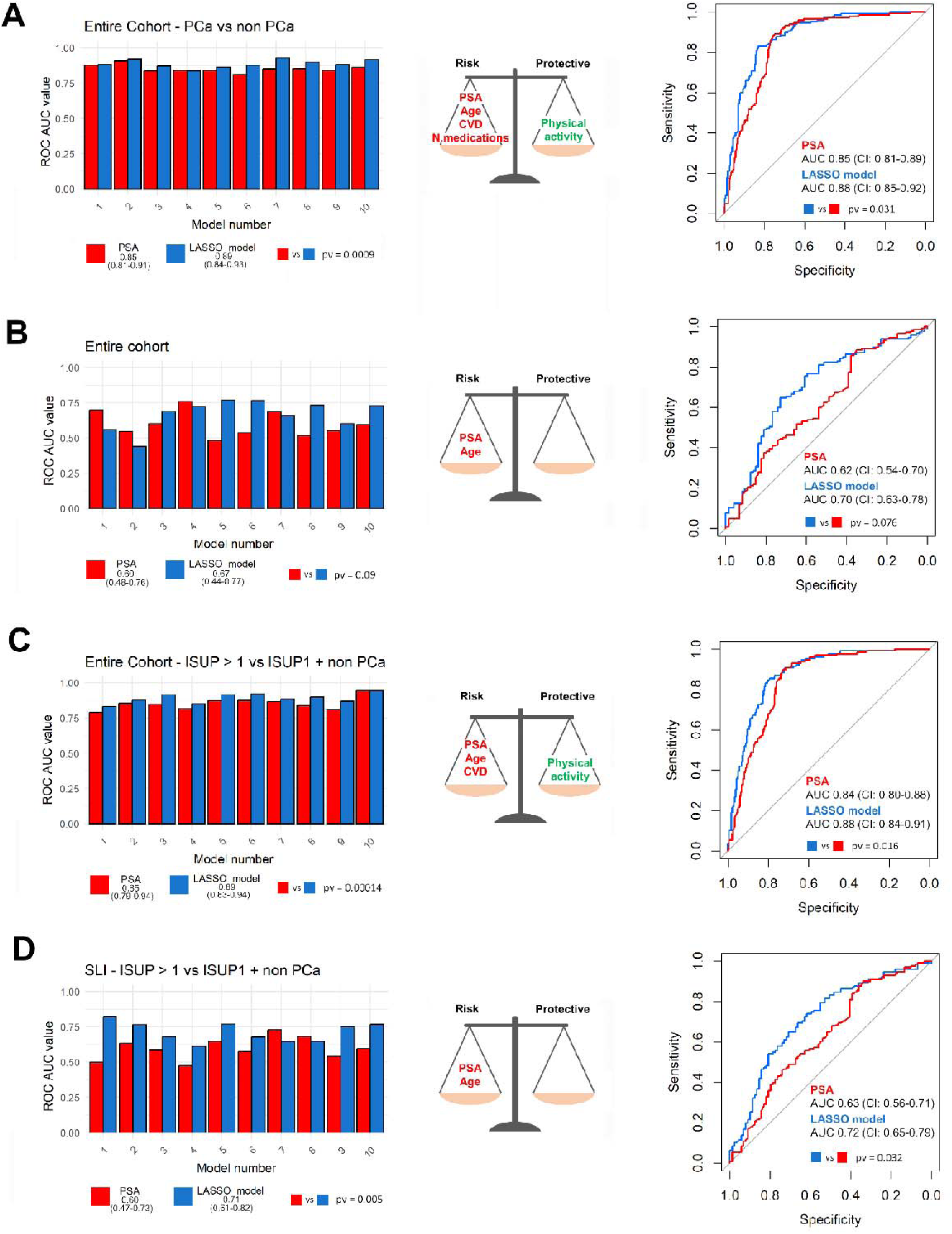
Left panels: histograms of the ROC AUCs for PSA (red) and the LASSO models tested on the 10 test sets (blue), with average AUCs on 10 test sets, ranges, and the p-values of the Student paired test between the 10 AUCs for PSA and the LASSO models. Central panels: risk and protective variables selected by 9/10 or 10/10 (bold) models. Right panels: ROC-AUCs for PSA (red) and the LASSO model (blue) on the test set. A refers to the entire cohort, with “non-PCa” as reference set; B refers to the group of men who underwent SLI, with “non-PCa” as reference set; C refers to the entire cohort, with ISUP1 PCa included into the reference set together with “nonPCa”; D refers to the group of men who underwent SLI, with ISUP1 PCa included into the reference set together with “nonPCa”.

When the same analysis on the entire cohort was conducted including ISUP1 PCa in the reference set, the LASSO models outperformed PSA alone (**Figure 3C, left panel**); PSA, age and CVD were selected by 10/10 models, hypertension by 9/10. When the outcome was high grade PCa (ISUP>2), PA was selected by 10/10 models, together with CVD, PSA and age, reaching an AUC of 0.86 (range: 0.76-0.91), compared to 0.81 (range: 0.73-0.89) of PSA alone (Student paired t-test p-value = 0.00026). In men who underwent SLI, the ROC AUCs were poor (**Figure 3D, left panel**) and no clinical variable except for CVD was selected to detect csPCa, indicating the need of stronger predictors.

As alternative approach, the initial group of 814 patients was divided in a training and a test set (**Figure 1**, step 1b), with similar proportions of cases (30% and 39%, respectively). LASSO regression was applied to the training sets (all the samples or SLI only), and the resulting models with selected variables (**Figure 3**, **middle panels**) were then applied to the corresponding test sets (**Figure 3, right panels**). As with the previous approach, PSA was outperfomed by models that include age, or age and anamnestic/lifestyle variables. However, especially when considering only men recalled for SLI, ROC-AUCs were not performant.

### Targeted sphingolipid analysis

Leveraging our previous findings on ceramide-like metabolites useful for the discrimination of PCa when PSA is in the so called grey-zone [3], we carried out targeted sphingolipid analysis on 369 samples available and previously used as test set (**Figure 1**, step 2). Fifty sphingolipids, including ceramides, dihydro-ceramides, hexosyl-ceramides, sphingomyelins, dihydro-sphingosine, sphingosine and its phosphorylated form, were analyzed on plasma samples using LC-MS/MS. All these molecules are characterized by an 18-carbon sphingoid base but harbor variable number of carbons in the fatty acids, ranging from 14 to 24. 46 sphingolipids were detected in all samples and were retained for the analysis.

LASSO regression modelling with 10-fold cross-validation was applied and the average ROC AUC of the 10 models applied to the test sets was 0.92 (range: 0.85-0.99), significantly higher compared to PSA alone and PSA combined with age, respectively (**Figure 4A, left panel**).

**Figure 4:**
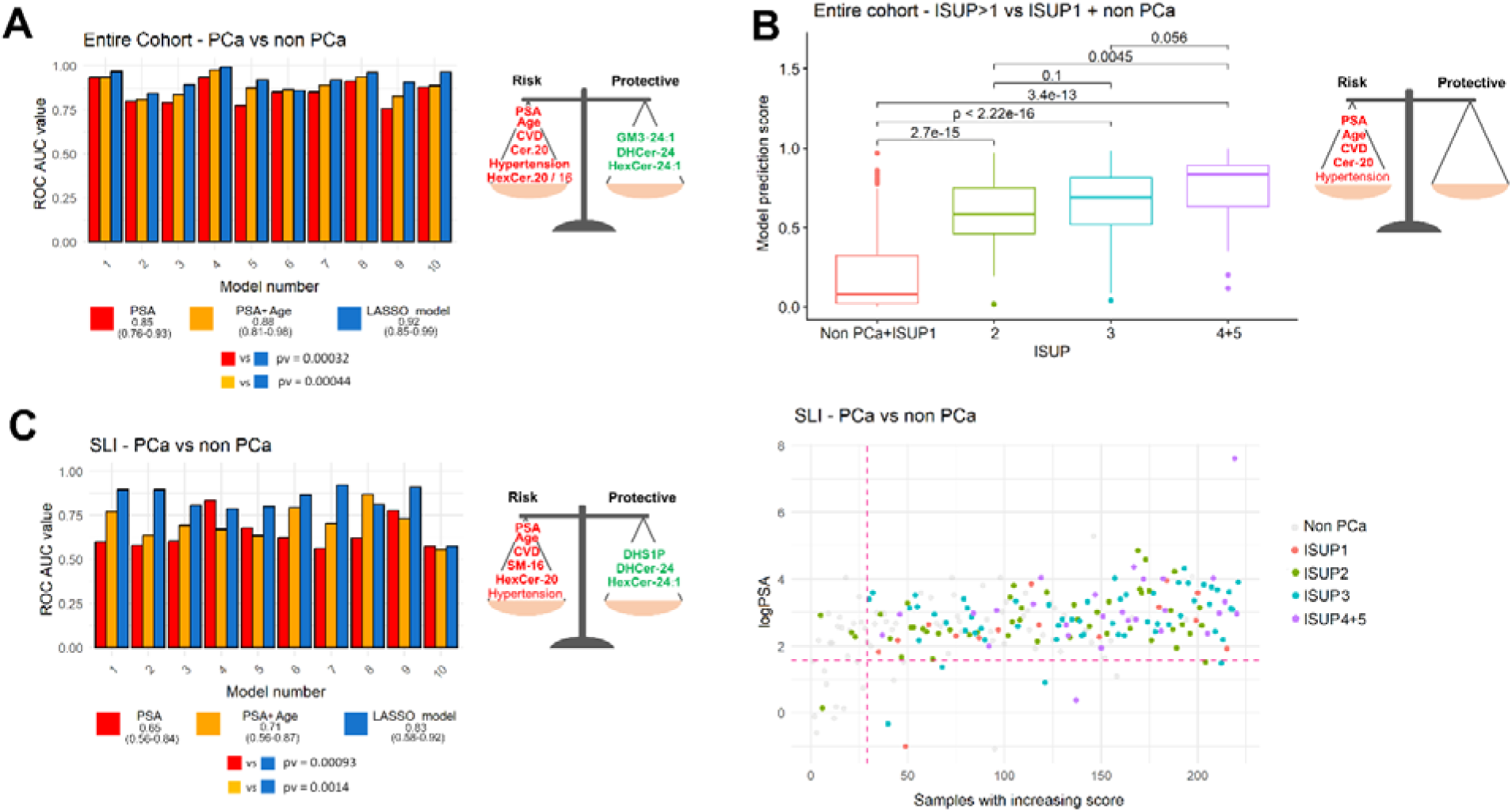
(A) Classification performances of the multivariable models on the 10 test sets vs PSA alone in the entire cohort with complete information on sphingolipids and lifestyle/clinical variables. (B) Predicted probabilities of harboring a PCa of ISUP grade >1 (csPCa) given by the multivariable model built using PSA, age, CVD and Cer-20. (C) Classification performances of the multivariable models vs PSA alone in the subgroup of men who underwent SLI (left panel); variables selected by 10/10 models (middle panel); representation of logPSA in men who were recalled for SLI, ordered by the predicted probabilities of harboring PCa using the variables shown in the middle panel (right panel). Grey dots correspond to nonPCa, red to ISUP1 PCa, green to ISUP2 PCa, light blue to ISUP3 PCa and violet to ISUP4-5 PCa. The horizontal dashed line corresponds to PSA = 3ng/ml, the vertical dashed line to the cut-off of 34% on the predicted probability.

Hypertension, CVD, PSA, age and five sphingolipids (HexCer-20 and Cer-20 with positive coefficients; HexCer-24.1, GM3-24.1, DHCer-24 with negative coefficient) were selected in 10/10 models (**Figure 4A, right panel**). **Figure 4B** shows the predicted probabilities of harboring csPCa given by the combination of the variables selected in 10/10 models when the predicted outcome was csPCa (PSA, age, CVD and Cer-20). For high grade ISUP >2 PCa, all 10 LASSO models identified a greater number of relevant variables, with four showing a positive association with the outcome (PSA, age, CVD, Cer-20) and eight showing a negative association (physical activity, SM-24, HexCer-24.1, GM3-18, LacCer-24, Cer-16, Gb3-24, DHCer-24). The mean AUC was 0.92 (range 0.80-0.98) for the 10 LASSO models, 0.81 (range 0.67-0.96) for PSA alone, and 0.83 (range 0.63-0.99) for PSA combined with age. The paired Student’s t-test for the AUCs across the ten models showed a statistically significant improvement over both PSA alone and PSA combined with age (p-value: 0.002 and 0.004, respectively).

The same approach was applied considering only the men who underwent SLI (n=221). In this case, CVD, PSA, age and five sphingolipids were selected by 10/10 models. Namely, SM-16 and HexCer-20 had positive coefficients, while HexCer-24:1, DHS1P, and DHCer-24 had negative coefficients (**Figure 4C, right panel**). The improvement in the average ROC AUC of the 10 models on the 10 test sets not used for model construction was even better, from 0.65 (range: 0.56-0.83) and 0.71 (range: 0.56-0.87) for PSA and PSA combined with age, respectively, to 0.83 (range: 0.58-0.92) for the model including CVD and sphingolipids (paired Student’s t-test p-value 0.0009 and 0.0014, respectively) (**Figure 4C, middle panel**). **Figure 4D, right panel** shows PSA values (log2 scale) in men who were recalled for SLI, ordered by the predicted probabilities of harboring PCa. Six ISUP>1 PCa (green, blue, and violet dots below the horizontal dashed line at PSA = 3□ng/mL and to the right of the model cut-off line), which would have been missed by PSA-based stratification alone, are correctly classified by the model. Moreover, three non-PCa (grey dots below the horizontal and to the right of the vertical dashed line) are misclassified only by the model (but not by PSA), whereas fourteen non-PCa (grey dots above the horizontal and to the left of the vertical dashed line) are misclassified only by PSA (but not by the model).

## Discussion

With the implementation of PCa screening policies across European countries, such as PRAISE-U [20], there will be an exponential rise in PSA testing. While this increase enhances early detection, it also raises the risk of dispensable second-level investigations (SLI), which can lead to over diagnosis, patient anxiety, and potential complications from invasive procedures like biopsies [21]. To optimize screening benefits and minimize harm, it is crucial to apply risk-stratified approaches and follow evidence-based guidelines to ensure that only high-risk individuals undergo further diagnostic assessments.

In this work, we summarized the results of a prospective multicentre study whose goal was to improve PSA diagnostic accuracy and reduce unnecessary MRIs/biopsies, by integrating lifestyle, age, comorbidity, and easily measured blood markers, ultimately informing targeted prevention strategies. A secondary endpoint was to accurately detect clinically significant PCa (ISUP>1) and high grade PCa (ISUP>2), in order to avoid overdiagnosis and overtreatment.

PSA dosage remains the milestone for PCa detection, even if with its limitations. However, the predictive value of PSA is drastically reduced in those men with PCa suspicion, candidates for SLI; in this subgroup of subjects, only about 60% really have a confirmed PCa diagnosis, actually making necessary the identification of other complementary biomarkers.

We found that the presence of cardiovascular diseases (CVD) increase the likelihood of a diagnosis of PCa of any grade, as well as of csPCa and high grade PCa. The association between CVD and prostate cancer should not be considered in the same way that smoking is a risk factor for lung cancer, as both CVD and PCa share some common risk factors, such as age, diet, obesity, lack of exercise, and levels of blood lipids (LDL, HDL, triglycerides) [22]. Nevertheless, a recent study highlighted a causative association between angina pectoris and PCa [23].

We highlighted a protective role of moderate and high-level physical activity (PA), and also highlighted that combining PA and CVD, along with PSA and age, improves patient selection for SLI. Notably, sedentariness is a modifiable risk factor that can influence the other variables. Our observations of higher PSA levels in sedentary men and decreased PSA levels proportional to exercise frequency in a subgroup of sedentary men that participated in a biweekly Nordic-walking program for one year within the DP3 study (manuscript in preparation), further underscores this point. Exercise impacts overall health, including cardiovascular health, and can even influence biological age [24]. These findings highlight the critical importance of stratifying patients for PCa screening and implementing risk-adapted strategies. By considering modifiable risk factors like sedentariness, alongside established factors such as PSA and age, we can more effectively identify individuals who would benefit most from further investigation, reducing unnecessary interventions in lower-intermediate risk individuals while ensuring timely diagnosis and treatment for those at higher risk. Critically, risk-adapted strategies could include the prescription of targeted exercise programs for sedentary men, addressing a key modifiable risk factor and potentially impacting both PSA levels and overall health.

The major finding of our work is that combination of risk factors with the analysis plasma sphingolipid levels further enhances the precision in detecting PCa, not only in the cohort including men without suspected PCa (average ROC AUC 0.92), but also among men recalled for suspected PCa (average ROC AUC 0.83). These results are comparable to risk calculators using MRI data [25] (AUC 0.81-0.87 for csPCa) and significantly better than models without MRI data [2] (AUC 0.66-0.79). While pre-biopsy MRI in suspected PCa can help over 30% of men avoid immediate biopsy and improve cancer detection – identifying about 15% more csPCa and 40% fewer clinically insignificant cancers [26] – it also increases the demand for MRI equipment, highly trained staff, manpower and AI-based image analysis. Using a pre-MRI risk assessment tool in a clinical cohort could eliminate over a third of prostate MRIs (or biopsies, if MRI is unavailable) [27], with even greater potential in screening populations where the overall risk of PCa is much lower. Therefore, MRI should be prioritized for men at intermediate and high risk based on a reliable stratification tool [28]. The drawback is a potential risk of missing some clinically significant PCas in men considered low-risk [27]. Nevertheless, our model with 5 sphingolipids combined with clinical/demographic covariates allowed the correct classification of six ISUP>1 PCa that would have been missed by a stratification based on PSA values only, with significant reduction of false-positives wihout missing any high grade PCa.

Interest in circulating sphingolipids as cancer biomarkers has grown rapidly in recent years due to their role as not just structural components of cell membranes, but also active molecules involved in various cellular processes, including cell growth, differentiation, and death. Most of the previous studies on sphingolipids and PCa regard patients with existing PCa diagnoses and have identified sphingolipid signatures associated with cancer progression and therapy resistance [7–12]. A prognostic blood-based sphingolipid panel has recently been proposed for men with localized PCa on active surveillance. This panel aims to identify patients at high risk of biopsy Gleason grade upgrading [29]. To our knowledge, no blood-based sphingolipid panel exists for the early detection of PCa within a screening context, nor as a pre-MRI/biopsy risk assessment tool for men with suspected PCa.

A limitation of this study is the incomplete availability of certain levels of information across enrolled subjects. We maximized the use of available data without imputing missing values. For multivariable modeling, especially when sphingolipids were included, we employed a 10-fold cross-validation approach to avoid overfitting and identify the most robust variables. In future work, we plan to expand our analyses to larger datasets and to integrate plasma proteomic data, to further validate our findings and provide a more comprehensive understanding.

This study investigated the potential of combining readily available information (PSA, age, hypertension, CVD, and sedentariness) with novel circulating biomarkers to improve PCa detection. Specific sphingolipid panels were identified that, when integrated with anamnestic/lifestyle factors, demonstrated strong diagnostic potential for both PCa screening and pre-MRI/biopsy risk stratification. These findings suggest that incorporating sphingolipid analysis could enhance early PCa detection and help reduce unnecessary invasive procedures.

## Data Availability

All data produced in the present study are available upon reasonable request to the authors

## Consent for publication

NA

## Availability of data and materials

Upon reasonable request to the corresponding author.

## Authors’ contributions

**CPN**: Conceptualization, Methodology, Investigation, Resources, Data Curation, Writing - Review & Editing. **PO**: Conceptualization, Software, Validation, Formal analysis, Data Curation, Writing - Review & Editing, Visualization. **MS**: Formal analysis, Investigation, Resources, Data Curation, Visualization. **MMG**: Conceptualization, Investigation, Resources, Writing - Review & Editing. **IG**: Investigation, Resources. **FG**: Software, Writing - Review & Editing. **FC**: Resources. **FM**: Resources. **MDC**: Investigation. **RP**: Supervision. **AS**: Resources. **FF**: Resources. **NT**: Resources. **MO**: Conceptualization, Resources, Writing - Review & Editing. **AZ**: Conceptualization, Resources. **CF**: Resources, Data Curation. **AG**: Resources. **AN**: Funding acquisition, Writing - Review & Editing. **LC**: Data Curation. **PG**: Resources, Supervision. **SZ**: Resources, Supervision. **GC**: conceptualization, methodology, Writing - Original Draft and Review & Editing, Supervision, Project Administration and Funding acquisition.

All authors read and approved the final manuscript

## Acknowledgements

We thank all patients who agreed to participate in the DP3 study, as well as all the nurses, physicians and students supporting sample collection and processing.

## Authors’ information (optional)

