## Supplemental file 1 for "Integrating anamnestic and lifestyle data with sphingolipid levels for risk-based prostate cancer screening"

**Additional file 1**: List of sphingolipids analysed.

| **CERAMIDES** | **GANGLIOSIDES GM3** |
| --- | --- |
| Cer.14 | GM3.16 |
| Cer.16 | GM3.18 |
| Cer.18:1 | GM3.18:1 |
| Cer.18 | GM3.20 |
| Cer.20 | GM3.22 |
| Cer.22 | GM3.24:1 |
| Cer.24:1 | GM3.24 |
| Cer.24 | **GLOBOTRIAOSYLCERAMIDES** |
| **DIHYDROCERAMIDES** | Gb3.16 |
| DHCer.16 | Gb3.18 |
| DHCer.18:1 | Gb3.18:1 |
| DHCer.18 | Gb3.20 |
| DHCer.24:1 | Gb3.22 |
| DHCer24 | Gb3.24 |
| **HEXOSYLCERAMIDES** | Gb3.24:1 |
| HexCer.16 | **SPHINGOSINES** |
| HexCer.18:1 | Sph |
| HexCer.18 | S1P |
| HexCer.20 | DHSph |
| HexCer.22 | DHS1P |
| HexCer.24:1 | **SPHINGOMYELINS** |
| HexCer.24 | SM.16 |
| **LACTOSYLCERAMIDES** | SM.18:1 |
| LacCer.16 | SM.18 |
| LacCer.18:1 | SM.24:1 |
| LacCer.18 | SM.24 |
| LacCer.20 |  |
| LacCer.22 |  |
| LacCer.24:1 |  |
| LacCer.24 |  |
