## Supplemental file 2 for "Integrating anamnestic and lifestyle data with sphingolipid levels for risk-based prostate cancer screening"

**Additional file 2**

*Lifestyle score calculations*

The reported weekly consumption frequencies (“never”, “once”, “2-3 times”, “more than 3 times”) for each food in the questionnaire were therefore assessed, in particular: “Pasta”, “Rice”, “White bread”, “Wholegrain bread”, “White meat”, “Red meat”, “Fish”, “Fried food”, “Cereals”, “Legumes”, “Fruit”, “Vegetables”, “Sweets”, “Eggs”, “Diary products”. To each food or combination of foods, a value between -1 and +1 was given according to the consumption frequencies recommended by the Italian Guidelines and the WCRF Recommendations. By adding the values attributed, an overall score for nutrition was calculated, ranging from 0 to 14.5, where higher scores indicated more adequate dietary habits, while lower scores an unhealthy diet. This score was further categorized into 5 classes: class 1 (0≤score≤7), class 2 (7<score≤8.75), class 3 (8.75<score≤10), class 4 (10<score≤11.5), class 5 (11.5<score≤14.5). Finally, the five scores thus created for BMI, physical activity, smoke, alcohol and diet were summed to obtain the final lifestyle score: higher lifestyle scores reflected a healthier global lifestyle, while lower scores were associated with worse lifestyle habits, predisposing to an increased risk of many pathologies.
